# Renovascular hypertension drives ischemic white matter injury through brain endothelial AT1R signaling

**DOI:** 10.64898/2026.10.15.26365689

**Authors:** Xuelian Jin, Hanki Kim, Hee Jin Jeong, Yuexian Cui, Shivani Shailendrakumar Singh, Hyo Gyeong Seo, Song E Kim, Hyang Woon Lee, Tracy D. Farr, Joanna M. Wardlaw, Simon R. Cox, Seong Hye Choi, Maria del C. Valdés Hernández, Jun Young Choi, Byung Gon Kim

## Abstract

**BACKGROUND:** White matter hyperintensities (WMHs), the principal imaging manifestation of cerebral small vessel disease, lack an animal model that produces pathology of sufficient magnitude to validate potential therapeutics. We characterized white matter (WM) degeneration in a renovascular hypertension (RVH) rat model, interrogated its mechanism, and tested its relevance in patients.

**METHODS:** RVH was induced by two-kidney, two-clip surgery and assessed by T2-weighted and diffusion MRI, myelin and axonal histology, and blood-brain barrier (BBB) leakage markers. The influence of angiotensin II (Ang II) on barrier function in brain endothelial cells was examined using transendothelial electrical resistance and reactive oxygen species (ROS) assays. Losartan was infused intracerebroventricularly to block brain AT1R (Ang II type 1 receptor) without systemic blood pressure reduction. WMH burden was compared between patients with and without renal artery stenosis, and WMH progression across antihypertensive classes was assessed in the CHALLENGE (Korea), MSS2 (UK), LBC1936 (UK), and SPRINT-MIND-MRI (US) cohorts.

**RESULTS:** Approximately two-thirds of RVH rats developed WMHs that expanded between 7 and 9 weeks, accompanied by reduced fractional anisotropy, demyelination, axonal and mature oligodendrocyte loss, and glial activation. IgG, albumin, and fibrinogen leakage was confined to affected WM. Ang II impaired endothelial barrier function and generated ROS through AT1R. Intracerebroventricular losartan attenuated WM pathology and improved behavior while blood pressure remained comparably elevated. Patients with renal artery stenosis had higher WMH volumes (10.52 versus 6.52 mL; *P* = 0.035). Angiotensin receptor blocker use was associated with the slowest tertile of annual WMH progression in the Korean cohort (OR, 2.57 [95% CI, 1.20–5.61]) and among White participants in SPRINT-MIND-MRI (OR, 3.52 [95% CI, 1.05–12.29]).

**CONCLUSIONS:** RVH produces prominent ischemic WM degeneration driven by AT1R signaling, identifying a blood pressure–independent therapeutic target with corroborating clinical evidence.

## Introduction

White matter hyperintensities (WMHs) constitute a principal imaging manifestation of cerebral small vessel disease (CSVD), a leading cause of cognitive impairment and neurological deficits in the aging population.^1, 2^ Vascular dementia, a severe form of vascular cognitive impairment, accounts for 15 to 20% of dementias from all causes in North America and Europe, and a higher proportion in Asian countries.^3^ Moreover, Alzheimer’s disease pathology frequently overlaps with vascular lesions, especially in the subcortical WM.^4, 5^ Targeting vascular contributions may slow the progression of degenerative dementia.^6^ Therefore, unraveling the mechanisms underlying ischemic WM injuries is warranted.

WMHs are accompanied by axonal loss, myelin pallor, impaired axon-glial junctions, blood-brain barrier (BBB) disruption, and glial activation.^7, 8^ An animal model that faithfully recapitulates these features is essential for defining how WM pathology contributes to cognitive decline, yet no consensus model exists.^9^ Vascular occlusive models based on carotid artery occlusion or stenosis are widely used, but the reported WM abnormalities have been inconsistent across studies,^8, 10–12^ leaving an unmet need for models that produce WM lesions of sufficient magnitude to validate potential therapeutics.^9^

This may be more attainable by incorporating underlying risk factors into an animal model. Because hypertension is the most important modifiable risk factor, several studies subjected spontaneously hypertensive Wistar-Kyoto rats to vascular occlusion, yielding variable results.^8, 13, 14^ By comparison, the renovascular hypertension (RVH) rat model has received less attention in the context of inducing ischemic WM injury. We were drawn to this model by recent reports describing WM lesions in RVH rats that were so prominent as to be unequivocally discernible even at low magnification.^15–17^ We hypothesized that RVH produces robust, well-demarcated WM lesions amenable to rigorous quantification, that this degeneration is mechanistically driven by angiotensin II (Ang II) signaling through the type 1 receptor (AT1R) at the BBB, and that the same mechanism contributes to WMH burden in humans. To test this, we systematically characterized WM degeneration in RVH rats, examined AT1R-mediated BBB impairment *in vitro* and *in vivo*, and, in independent human cohorts, assessed WMH burden in patients with renal artery stenosis (RAS) and the longitudinal course of WMHs across antihypertensive classes including Ang II receptor blockers (ARBs).

## Materials and methods

### Animals

Male Sprague-Dawley rats at four weeks of age (weighing 70-90 grams) were purchased from Orient Bio Inc. The animals were housed in the Animal Experiment Center at Ajou University School of Medicine on a 12 h light/dark cycle with ad libitum access to food and water. All animal procedures were approved by the Ajou University School of Medicine Institutional Animal Care and Use Committee (AJOU-IACUC-2024-0028).

### Surgical procedures for clipping bilateral renal arteries

To establish the RVH rat model, we adopted the two-kidney, two-clip (2K2C) method as described in previous studies (Fig. 1A).^17, 18^ Briefly, four-week-old rats were anesthetized with 3% isoflurane and maintained at 2% isoflurane in a 30% oxygen and 70% nitrogen mixture. A midline longitudinal abdominal incision was made, and the small intestine was gently retracted to expose the left and right renal arteries. A ring-shaped silver clip measuring 4 mm in length and 0.3 mm in diameter (generously donated by Prof. Yuhua Fan at Sun Yat-sen University, China) was placed and secured with hemostatic forceps. Animals in the sham-operated group underwent the same procedures without clipping. For initial characterization of the RVH rat model, 32 animals underwent 2K2C surgery, while 8 animals underwent sham surgery. It was notable that 8 out of 32 (25%) died during the follow-up period, suggesting that these animals may have developed 2K2C-related severe lesions.

**Figure 1.**
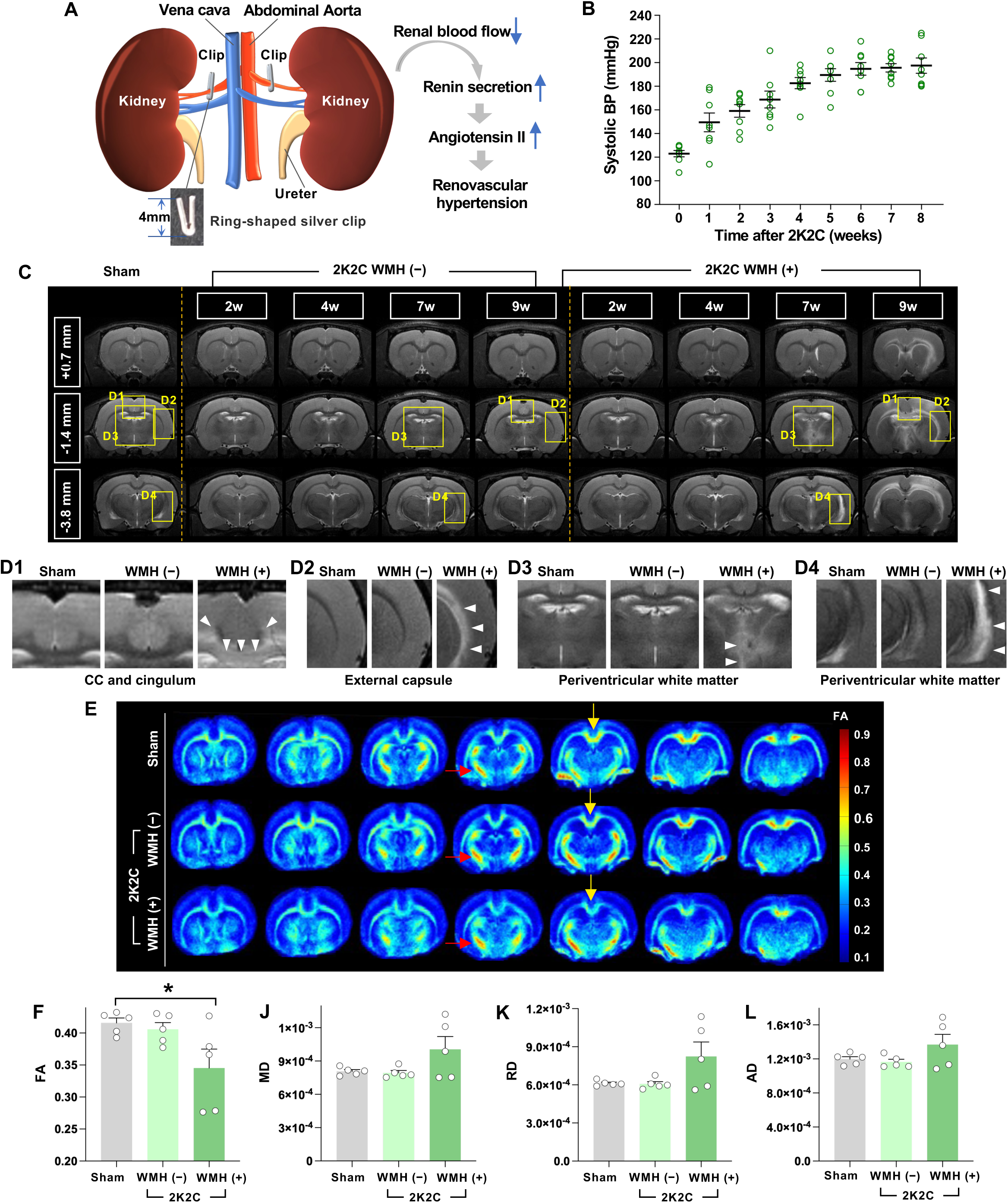
Renovascular hypertension induces progressive white matter hyperintensities in rats. **(A)** Illustration of the renovascular hypertension (RVH) model induced by the two-kidney, two-clip (2K2C) procedure. A photo of a silver clip is shown. Reduced renal blood flow induces renin secretion, leading to increased angiotensin II and, finally, RVH. **(B)** Changes in systolic blood pressure (SBP) are shown in 8 animals with serial BP measurements obtained until 8 weeks after 2K2C surgery. **(C)** Detection of white matter hyperintensities (WMHs) with T2W MRI following the 2K2C procedure. Representative coronal T2W MR images at different time points from the same animal in each group. Images were obtained at 0.7 mm rostral, 1.4 mm, and 3.8 mm caudal to the bregma. WMHs were observed in the 2K2C WMH (+) group at 7 and 9 weeks. **(D)** Enlarged MR images from the yellow rectangles labeled D1-4 in **(C)** are shown below. White arrowheads indicate WMHs in the corpus callosum (CC) and cingulum **(D1)**, external capsule **(D2)**, and periventricular white matter surrounding the third ventricle **(D3)** and lateral ventricle **(D4)**. **(E)** A color-coded fractional anisotropy (FA) map calculated using diffusion MRI (dMRI) from each representative animal from each group. Yellow arrows indicate decreased FA in the CC in the 2K2C WMH (+) group, and red arrows indicate no change in FA in the internal capsule. **(F)** Quantification graphs comparing the FA, mean diffusivity (MD), radial diffusivity (RD), and axial diffusivity (AD) in the CC among sham, 2K2C rats with and without WMH lesions. and the control rats. * indicates *P* < 0.05, by one-way ANOVA followed by Tukey’s *post hoc* analysis. N = 5 animals per group. Error bars represent the SEM.

### Blood pressure (BP) measurement

BP was measured using a non-invasive tail-cuff BP monitoring system with 4 active channels (CODA^®^ High Throughput System, #CODA-HT4, Kent Scientific).^19^ Each recording session consisted of 20 expansion and contraction cycles; the first 5 were “adaptation” cycles not included in the analysis, and the following 15 were considered “regular” and included in the final measurements. After excluding values the system marked as false, we calculated the average systolic BP (SBP) from at least 5 readings. If a session had fewer than 5 valid measurements, or if the standard deviation of the 5 measured values exceeded 30 mmHg, the entire session of 20 cycles was repeated.

### Magnetic resonance imaging (MRI)

A subset of 20 rats, including 5 sham-operated controls, underwent serial MRI. Two rats in the 2K2C group died during follow-up and were therefore excluded from the analysis (final n for 2K2C group = 13). MRI was performed on a 9.4 T/30 cm Bruker BioSpec MR system. Rats were anesthetized, and respiratory rate and body temperature were monitored using a Model 1030 (Small Animal Instruments Inc.). A T2-weighted (T2W) 2D Turbo Rapid Acquisition with Relaxation Enhancement (RARE) sequence was acquired: TR/TE = 3200/32 ms, resolution = 117 μm^2^; number of 800 μm slices = 30, RARE factor = 8, averages = 5, scan time = 10 min 16 sec.

Diffusion MRI (dMRI) was conducted on 5 sham controls and 10 of the 13 animals in the 2K2C group after the final structural MRI session, including 5 with WMH and 5 without WMH lesions. A spin-echo sequence with the following parameters was used: TR/TE = 3000/35 ms, field of view = 30 × 15 mm^2^; matrix = 256 × 128, in-plane resolution = 0.117 mm^2^, diffusion encoding gradient directions = 30, gradient duration (δ) = 2.5 ms, gradient separation (Δ) = 8.5 ms, and b-value = 1000 s/mm^2^. Diffusion scalar images were processed using the diffusion toolbox in FMRIB’s Software Library (FSL). Non-brain tissue was removed using DTI Studio. For each animal, the skull-stripped b0 images were averaged to generate a mean b0 image, which was then manually coregistered to the corresponding individual T2W anatomical image using MRIcro (nitrc.org/projects/mricro/). Coregistration was performed separately for each animal by visually aligning corresponding anatomical landmarks between the mean b0 image and the individual T2W anatomical image. The inverse of the resulting b0-to-T2 transformation was applied to map the anatomically defined regions of interest (ROIs) from the T2W anatomical space into the native diffusion space. Anatomical localization of the ROIs was guided by the Small Animal Molecular Imaging Toolbox (SAMIT) (https://mic-umcg.github.io/samit/), which was developed based on the Paxinos anatomical atlas.^20^ ROIs were manually delineated on each animal’s individual T2W anatomical image using MRIcro, with anatomical localization guided by the SAMIT rat brain atlas. The transformed ROIs were applied to the corresponding fractional anisotropy (FA), mean diffusivity (MD), radial diffusivity (RD), and axial diffusivity (AD) maps. We extracted mean FA, MD, RD, and AD values from each ROI for quantitative analysis.

### Intracerebroventricular infusion of the AT1R antagonist

To examine the impact of AT1R on WMH, an additional 52 rats underwent sham surgery (n = 18) or 2K2C surgery (n = 34). Four weeks later, they were further randomized to receive intracerebroventricular (ICV) injections of vehicle (sham, 9; 2K2C, 18) or losartan (sham, 9; 2K2C, 16). A total of 11 rats died during the follow-up period and thus were excluded from the final behavioral and histological analyses. The Brain Infusion Kit 2 (Alzet) was connected to an osmotic minipump (model 2004). Animals received intraperitoneal injections of ketamine-xylazine (80 mg/kg and 10 mg/kg), were placed in a stereotaxic frame, and had cannulas implanted in the right ventricle using the following coordinates: 0.8 mm posterior, 1.6 mm lateral, and 5 mm deep. The cannula was secured with adhesive gel and the minipump implanted subcutaneously on the dorsal side. Pumps contained either the vehicle artificial CSF (aCSF) or losartan (12.5 μg/h) dissolved in aCSF. Solutions were delivered continuously at 0.25 μl/h for 28 days.

### Quantitative image analysis

Imaging parameters, including gain, digital offset, and laser intensity, were held constant. Images were analyzed using ImageJ (US National Institutes of Health, https://imagej.nih.gov/). To characterize the extent of WM degeneration in 2K2C rats, three coronal sections spaced 1680 μm apart were selected, and 0.1 mm^2^ ROIs were placed at the center of the cingulum. The severity of WM lesions in ECRC staining was graded by a blinded experimenter (HGS): Grade 0 normal, Grade 1 focal decrease in stain intensity (rarefaction) limited to the cingulum, Grade 2 diffuse decrease in the density of myelin staining in the cingulum and other parts of the corpus callosum (CC) in one of three sections, and Grade 3 diffuse rarefaction in at least two of the three sections. The average MBP immunoreactivity intensity was measured and divided by the average intensity in the sham group to calculate the relative fold change. To quantify axonal degeneration, the area fraction of NF-M immunoreactive signals above a preset threshold was measured in ImageJ. GFAP and Iba-1 immunoreactivities were measured to compare the extent of astrocytic activation and microglial inflammation, respectively. For quantitative comparison of cell numbers, APC-CC1- and NG2-positive cells with clear DAPI nuclear signals were counted and expressed as cell density per 0.1 mm^2^.

### Analysis of WMH burden on MRI in a retrospective cohort of human patients with confirmed renal artery stenosis (RAS)

This retrospective study was approved by the Ajou University Hospital Institutional Review Board (AJOUIRB-MDB-2022-158). Among 210 patients screened between 2002 and 2021 at Ajou University Hospital, South Korea, who underwent renal artery computed tomography (CT) for suspected secondary hypertension and subsequent brain MRI, 109 were excluded due to end-stage renal disease, kidney transplantation, renal thrombus, acute stroke, intracranial hemorrhage, brain tumors, or poor scan quality. For serial scans, the MRI taken closest to the renal artery CT scan date was analyzed. Total WMH volume was segmented from T2-fluid-attenuated inversion recovery (FLAIR) images using the lesion prediction algorithm in the Lesion Segmentation Tool (LST v3.0.0) for SPM12.^21^ Clinical data were extracted from electronic medical records and summarized in Supplementary Table 1.

### *Post hoc* analyses of the potential influence of various antihypertensive medications on WMH in clinical cohorts

To evaluate class-specific associations of antihypertensive medications with WMH burden progression, we conducted *post hoc* analyses of four independent clinical cohorts from South Korea, the UK, and the US, encompassing populations of diverse ancestry. Demographic information and baseline clinical characteristics of the four cohorts were summarized in Supplementary Tables 2−5. The CHALLENGE study is a multicenter, double-blind, randomized clinical trial (RCT) in South Korea that evaluates cilostazol versus aspirin in patients with CSVD.^22^ Among 256 subjects initially randomized, 189 who completed the 2-year follow-up MRI scan were finally included in the analysis. The current retrospective *post hoc* clinical study protocol was approved by the Institutional Review Board (IRB) of Ajou University Hospital (AJOUIRB-DB-2025-302). The Mild Stroke Study 2 (MSS2) is a prospective observational cohort of patients in Scotland, UK, presenting with symptoms of lacunar or mild cortical ischemic stroke.^23^ Of the 264 initially recruited patients, only 191 with complete 1-year follow-up MRI data were included. The MSS2 was approved by the Lothian Ethics of Medical Research Committee (REC 09/81,101/54) and the NHS Lothian R+D Office (2009/W/NEU/14). The Lothian Birth Cohort 1936 Study (LBC1936) is a longitudinal observational study of factors that influence cognitive aging, comprising Scottish participants from the Scottish Mental Health Survey of 1947 and including a 3-year follow-up MRI.^24^ We used the following data from the LBC1936 database (https://lothian-birth-cohorts.ed.ac.uk/data-access-collaboration [Accessed on 16/04/2026]): age at the time of the first brain MRI scan (study testing wave 2), sex, white matter hyperintensity (WMH) volume in mm^3^ from the first and second brain scans, WMH volume as percentage in the intracranial volume, and white matter damage also calculated from the first two brain MRI scanning sessions (i.e., testing waves 2 and 3), intracranial volume, medications at waves 2 and 3, and the following self-reported information: hypertension (y/n), diabetes (y/n), and hypercholesterolaemia (y/n) from wave 2, 465 patients who completed the second and third wave of the study, with fully intact MRI data, were included in the *post hoc* analysis. The LBC1936 was approved by the Multi-Centre Research Ethics Committee for Scotland (MREC/01/0/56) and the Lothian Research Ethics Committee (LREC/2003/2/29). All primary trials obtained written informed consent from participants. The SPRINT-MIND-MRI study is an RCT substudy in the US investigating the effect of intensive versus standard BP control on cerebral WMH.^25^ Using publicly available datasets from the NHLBI BioLINCC repository (https://biolincc.nhlbi.nih.gov/studies/sprint/), we retrospectively reassessed the influence of antihypertensive classes on WMH progression in a race-stratified manner, with race self-reported in the parent trial (AJOU-IRB-EX-2022-559). Of the 1267 randomized participants, 663 underwent baseline MRI, and 445 of these had a 4-year follow-up scan with complete clinical data. Ten individuals who self-identified as neither White nor Black were excluded because of the small numbers in these categories (final n = 435).

Across all the *post hoc* analyses, antihypertensive medications were categorized into six major classes: angiotensin-converting enzyme inhibitors (ACEis), ARBs, beta-blockers (BB), calcium channel blockers (CCBs), diuretics (DIUs), and other antihypertensives (OTHs). Medication exposure was defined as adherence to >70% of the follow-up period for the CHALLENGE and SPRINT-MIND-MRI studies, as baseline exposure in the MSS2, and as exposure at both the baseline and follow-up period in the LBC1936 study. The relative prescription frequency of the two antihypertensive medications targeting the renin-angiotensin system, ACEis and ARBs, differed markedly between the CHALLENGE study and the other cohorts. In the CHALLENGE Korean cohort, only 2 subjects received ACEi prescriptions, so we excluded ACEi use from multivariable modeling. In contrast, ACEi use was more prevalent than ARB use in the UK and US cohorts. We were concerned that the small number of ARB users might undermine the statistical power for the multivariable analysis of ARB medication and WMH progression. In addition to the analysis of individual medications, we therefore combined users of either class to test whether renin-angiotensin system inhibition as a whole was associated with WMH progression. WMH volumes were expressed as a percentage of corresponding intracranial volume. Annualized WMH progression was calculated as the difference between follow-up and baseline WMH volumes divided by the follow-up period (in years). The primary outcome was defined as annualized progression ≤ the 1st (slowest) tertile. Secondary outcomes included progression ≤ 0 (stable/regression) and ≤ the median (for the SPRINT-MIND-MRI study, both the total cohort median and the intensive-arm median were included as secondary outcomes). Multivariable logistic regression models were fitted to estimate odds ratios (ORs) and 95% confidence intervals (CIs) for antihypertensive use by class to assess the primary and secondary outcomes. Covariates included age, sex, baseline WMH volume, mean on-treatment SBP (in the MSS2 and LBC1936 study, self-reported hypertension diagnosis was used instead), trial randomization arm where applicable, diabetes mellitus, dyslipidemia, and the concurrent use of all other antihypertensive drug classes (mutually adjusted to evaluate class-specific independent effects).

### Statistical analysis

All numerical values and error bars in the quantification graphs are expressed as mean ± standard error of the mean (SEM). An unpaired Student’s t-test (two-tailed) was used to compare mean values between two groups. One-way ANOVA followed by Tukey’s *post hoc* analysis was used to compare the means of three or more groups. The statistical significance of the correlation between SBP and pathological grading in animals subjected to 2K2C surgery was assessed using a nonparametric Spearman correlation analysis. For weekly comparisons of neurological functions, a repeated-measures two-way ANOVA was conducted, followed by Bonferroni *post hoc* tests to assess group differences at each time point. Two-way ANOVA followed by Bonferroni’s post hoc analysis was performed in experiments with two independent variables. For clinical studies, group comparisons were performed using R (version 4.4.1) with two-tailed Student’s t-test for normally distributed continuous variables, the Mann–Whitney U test for non-normally distributed continuous variables, and the chi-square test (or Fisher’s exact test, as appropriate) for categorical variables. Quantification graphs were generated using GraphPad Prism (v11.0.0).

More detailed methods are available in the Supplementary Methods.

## Results

### RVH induces progressive white matter hyperintensities in rats

SBP began to rise 1 week after the 2K2C procedure, increased progressively through 6 weeks, then plateaued, with the mean SBP reaching approximately 200 mmHg (Fig. 1B). T2W MRI revealed unequivocal high signal intensity lesions in 8 out of 13 RVH animals (61.5%) at 9 weeks. The hyperintensities were predominantly located in and around the WM, including the CC, cingulum, and external capsule, but not the internal capsule (Fig. 1C, D). WMHs were also observed in the periventricular WM (Fig. 1D). Hyperintensity signals were not typically observed in the cerebral cortex and deep gray regions, such as the basal ganglia and thalamus (Fig. 1C, D). Intriguingly, 38.5% (5 of 13) of rats that underwent the same 2K2C procedure showed no apparent WM abnormalities (Fig. 1C). WMH appeared to develop between 4 and 7 weeks after 2K2C, with most signals typically appearing at 7 weeks. Importantly, WMH extent increased between the 7- and 9-week time points, indicating progressive evolution.

We performed dMRI to evaluate WM integrity. Fractional anisotropy (FA) appeared to decrease in the CC in the 2K2C WMH (+) group but not in the internal capsule (Fig. 1E). Mean FA in the CC differed significantly among sham, WMH (-), and WMH (+) groups (*F_(2, 12)_* = 4.203, *P* < 0.05), with the significant difference only between the sham and WMH (+) group by *post hoc* analysis (Fig. 1F). MD tended to increase only in the CC of rats with WMH lesions, but this was not significant (*F_(2, 12)_* = 2.978, *P* = 0.0891) (Fig. 1E). Similarly, RD showed a clear trend toward an increase only in the CC of rats exhibiting WMH (*F_(2, 12)_* = 3.358, *P* = 0.0695). AD did not differ significantly (Fig. 1E). There was no noticeable difference in the internal capsule among the three groups for FA, MD, RD, or AD (Supplementary Fig. 1).

### Pathological degenerative changes in the WM induced by RVH

Next, we evaluated the histopathological changes in RVH rats. The WM abnormalities were discernible in the brain tissue sections stained with ECRC even at a low magnification: the density of the myelin staining was noticeably decreased in the cingulum and the CC below the cingulum (Fig. 2A). In higher magnification images, the genu portion of the CC also revealed a decrease in the intensity of the myelin staining. White matter tracts in the external capsule also frequently showed focal or diffuse hypomyelination (Fig. 2B). We also examined the morphology of the blood vessels located near the white matter tract. Intimal thickening with luminal narrowing or obliteration was observed only in rats undergoing the 2K2C procedure (Fig. 2C). However, as in the MRI evaluation, not all animals exhibited degenerative changes in the WM; some rats subjected to the same 2K2C procedure showed no obvious pathological abnormalities (Supplementary Fig. 2). To semi-quantitatively assess the severity of RVH-induced white matter lesions, we developed a pathological grading scheme (Fig. 2D). Grade 0 accounted for 33.3%, Grade 1 for 8.3%, Grade 2 for 16.7%, and Grade 3 for 41.7% (Fig. 2E). There was a tendency for animals with severe lesions to have higher SBP, but this correlation was not significant (Fig. 2F).

**Figure 2.**
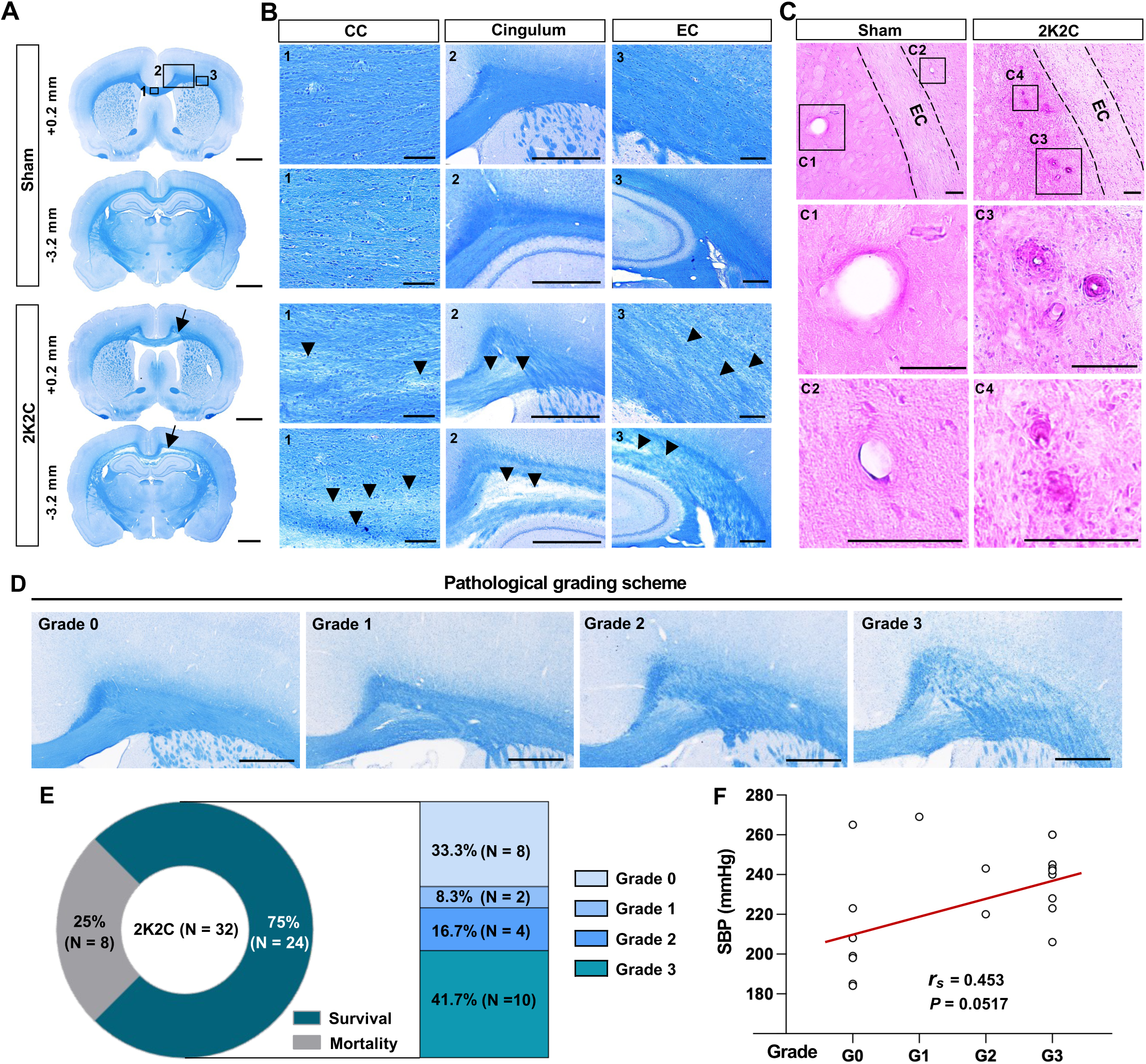
Pathological degenerative changes in the WM by RVH. **(A)** Histological assessment of the WM degeneration in RVH rats. Representative images of coronal brain sections stained with Eriochrome Cyanine RC (ECRC) to visualize the myelinated WM. Two sections at 0.2 mm rostral and 3.2 mm caudal to the bregma in the sham and 2K2C groups with WM lesions are shown. Black arrows indicate regions with decreased myelin density that are unequivocally notable in these low-magnification images in the 2K2C group. Rectangles labeled 1-3 indicate the regions from which the magnified images shown in **(B)** were obtained. Scale bars = 2 mm. **(B)** Higher-magnification images from ECRC-stained brain sections showing the corpus callosum (CC), cingulum, and external capsule (EC). Black arrowheads indicate WM lesions with focal or diffuse demyelination in these WM tracts. Scale bars = 200 μm. **(C)** H&E staining of small vessels in sham-operated and RVH rats. Intimal thickening with luminal narrowing or obliteration was observed in rats subjected to the 2K2C procedure. Rectangles labeled C1-4 represent regions that are magnified below in C1-4. Scale bars = 100 μm. **(D)** Semi-quantitative pathological grading scheme to assess the severity of WM degeneration: Grade 0 = no unequivocal lesion; Grade 1 = focal demyelination; Grade 2 = diffuse demyelination in only one section; and Grade 3 = diffuse demyelination in at least two of three sections. Scale bars = 500 μm. **(E)** A pie chart shows the mortality and survival rates of animals subjected to 2K2K procedures. Surviving animals are classified by pathological grade, including Grade 0, which indicates no unequivocal lesion. **(F)** Correlation between pathological grade and systolic blood pressure (SBP) measured at the last time point. *r_s_* = Spearman’s coefficient.

We subsequently divided RVH rats into those without (Grade 0) and those with WM lesions (Grade 1-3) to compare MBP immunoreactivity. There was no appreciable difference in Grade 0 rats compared to the sham surgery group, but the overall intensity was decreased in rats with apparent WM lesions (Fig. 3A, C). Intriguingly, the extent of demyelination appeared greater in posterior regions (Fig. 3A, C). Neurofilament staining also revealed decreased axonal density in the CC, especially in the more posterior CC (Fig. 3B, D). We also quantified changes in the number of mature oligodendrocytes in the CC between sham-operated and rats with WM lesions (Grade 1-3). The number of APC-CC1-positive cells colocalized with nuclear Olig2 signal was significantly decreased in animals with 2K2C-induced WM damage (Fig. 3E, G). In contrast, NG2-positive oligodendrocyte progenitor cells increased in these animals compared with sham-operated animals (Fig. 3F, H). Microglial activation in the CC was markedly increased in Grades 1-3 rats (Supplementary Fig. 3A, C), accompanied by astrocytic activation (Supplementary Fig. 3B, D), indicating that WM injury induced prominent neuroinflammatory reactions in the CC.

**Figure 3.**
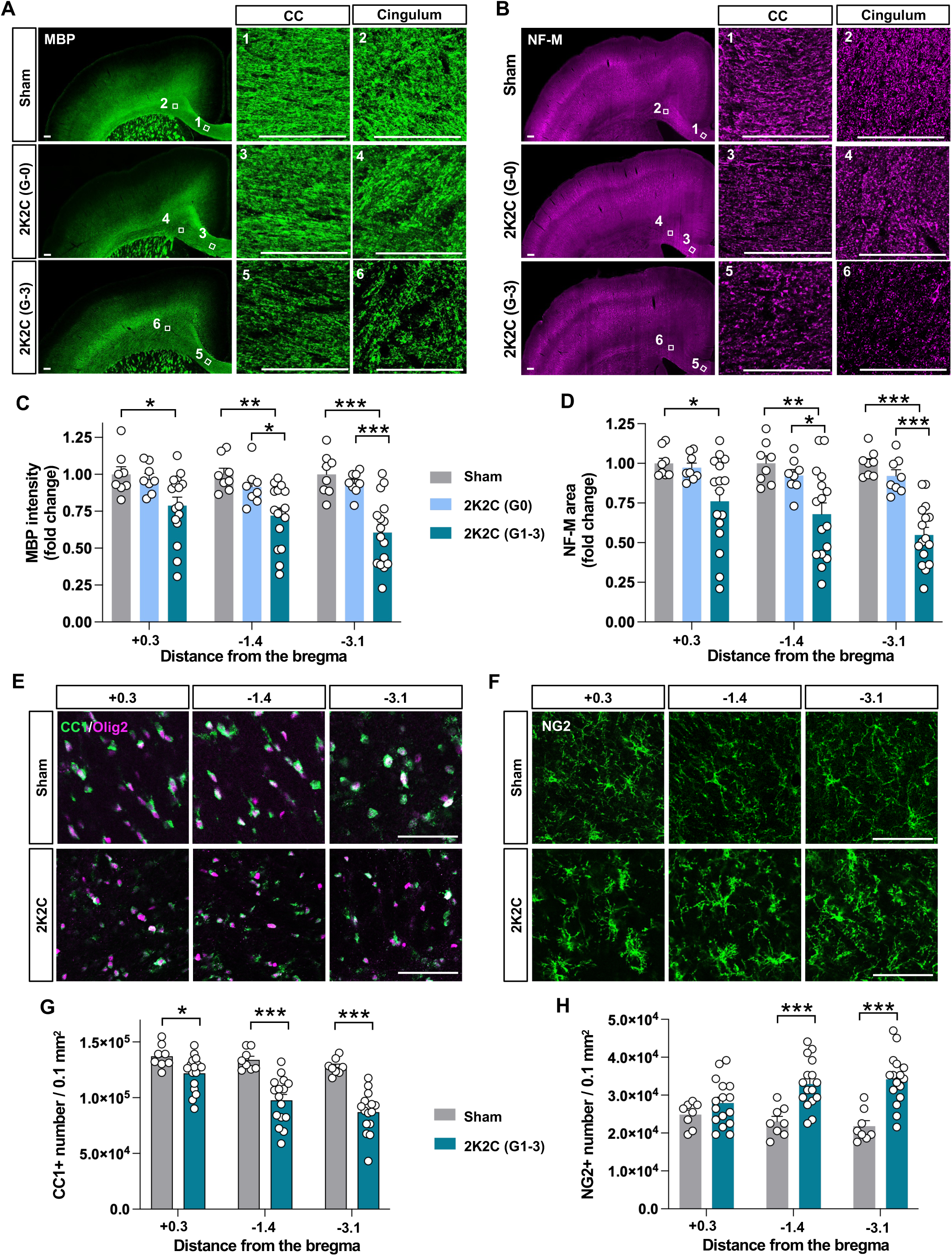
Quantitative evaluation of the demyelinating and axonal pathology in RVH rats. **(A-D)** Assessment of demyelination and axonal degeneration. **(A, B)** Representative images of coronal brain sections stained with antibodies against myelin basic protein (MBP) **(A)** and neurofilament-M (NF-M) **(B).** The boxed regions labeled with numbers are magnified on the right (1, 3, 5, CC; 2, 4, 6, cingulum). **(C, D)** Quantification graphs comparing the intensity of MBP immunoreactivity **(C)** and the area of NF-M positive axons **(D)**. Error bars represent mean ± SEM. *, **, and *** indicate *P* < 0.05, *P* < 0.01, and *P* < 0.001, respectively. n = 8 (sham), 8 (2K2C Grade 0), and 16 (2K2C Grade 1-3). Scale bars = 100 μm. **(E-H)** Assessment of oligodendrocyte lineage cells. **(E, F)** Representative images of coronal brain sections stained with both APC-CC1 and Olig2 antibodies for mature oligodendrocytes **(E)** and with NG2 antibodies for immature oligodendrocytes **(F)** in the cingulum. Tissue sections were selected at 0.3 mm rostral, 1.4 mm, and 3.1 mm caudal to the bregma. **(G, H)** Quantification graphs comparing the number of mature oligodendrocytes (CC1+ and Olig2+) **(G)** and NG2+ oligodendrocyte precursor cells **(H)**. Error bars represent mean ± SEM. *, **, and *** indicate *P* < 0.05 and *P* < 0.001, respectively. n = 8 (sham) and 16 (2K2C Grade 1-3). Scale bars = 50 μm.

### Association between RAS and WMH burden in humans

The striking pathological finding in the WM of the rats with RVH induced by the 2K2C procedure led us to explore its clinical relevance in human patients with RAS. Of the 210 patients who underwent renal artery CT angiography to evaluate secondary hypertension, 109 were selected after applying the exclusion criteria described in the Methods section (Fig. 4A). Ultimately, we compared 34 patients with RAS confirmed by CT angiography (Fig. 4B) with 34 controls propensity score-matched for age and antiplatelet use. Baseline characteristics did not differ significantly between the two groups, including age and BP at the time of the CT scan (Supplementary Table 1). More than 90% of subjects with and without RAS were receiving antiplatelet therapy. The percentage of subjects taking antihypertensive medications, including ACEis and ARBs, did not differ significantly between the two groups (Supplementary Table 1). After removing extreme outliers from both groups, the intergroup comparison showed significantly higher WMH volume in patients with RAS than in those without RAS (6.52 mL vs. 10.52 mL, P = 0.035, Mann–Whitney U test) (Fig. 4B, C).

**Figure 4.**
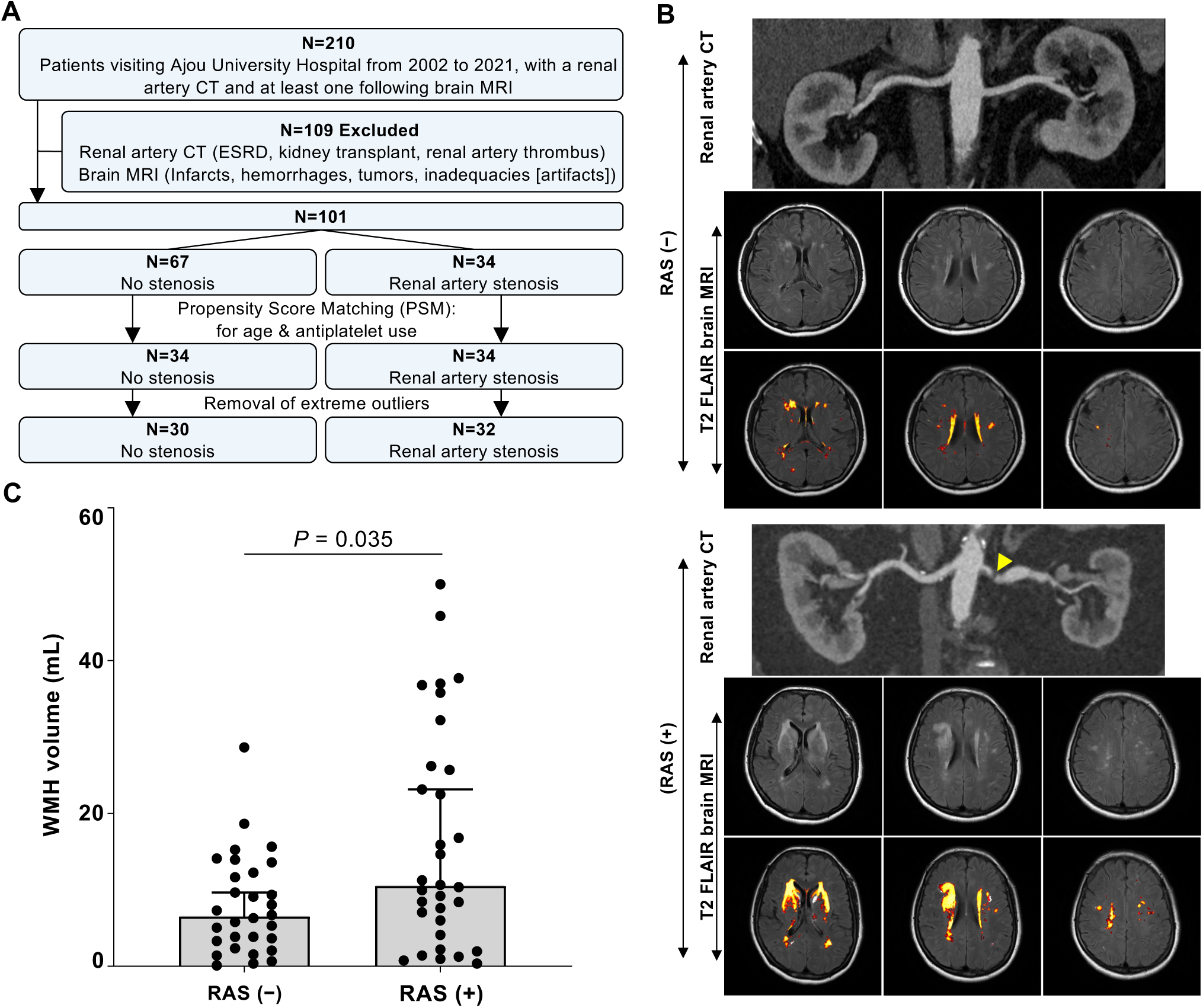
Association between renal artery stenosis and WMHs in humans. **(A)** Flowchart illustrating patient selection in the renal artery stenosis (RAS) cohort. (**B)** Representative renal artery CT angiograms and brain T2 fluid-attenuated inversion recovery (FLAIR) MRI images, with corresponding lesion maps segmented using the Lesion Segmentation Tool (LST) toolbox displayed across three representative brain levels. The yellow arrowhead highlights renal artery narrowing. (**C)** Quantification and comparison of WMH volume (mL) in patients with RAS (n = 32) and without RAS (n = 30) (data presented as median with 95% CI range, Mann-Whitney U test, two-tailed).

### Implication of Ang II/AT1R signaling in BBB impairment

There is mounting evidence that BBB impairment precedes WMH associated with hypertension.^26^ Indeed, tight junction proteins are also disrupted in RVH rats.^27^ We observed no albumin leakage in sham-operated rats, but very intense accumulation was observed in 2K2C rats exhibiting obvious WM pathology (Grade 1-3). Strikingly, albumin leakage was largely confined to the CC and cingulum, where WM pathology was observed, but was very rare in the cerebral cortex or other subcortical gray matter regions. (Fig. 5A). IgG leakage also showed a very similar pattern (Fig. 5A), and this was confirmed by quantification (Fig. 5B). To further substantiate these findings, we examined fibrinogen extravasation, another indicator of BBB disruption.^28^ We frequently observed fibrinogen immunoreactivity encircling the RECA-1-positive vessel walls in the cingulum in RVH rats (Fig. 5C).

**Figure 5.**
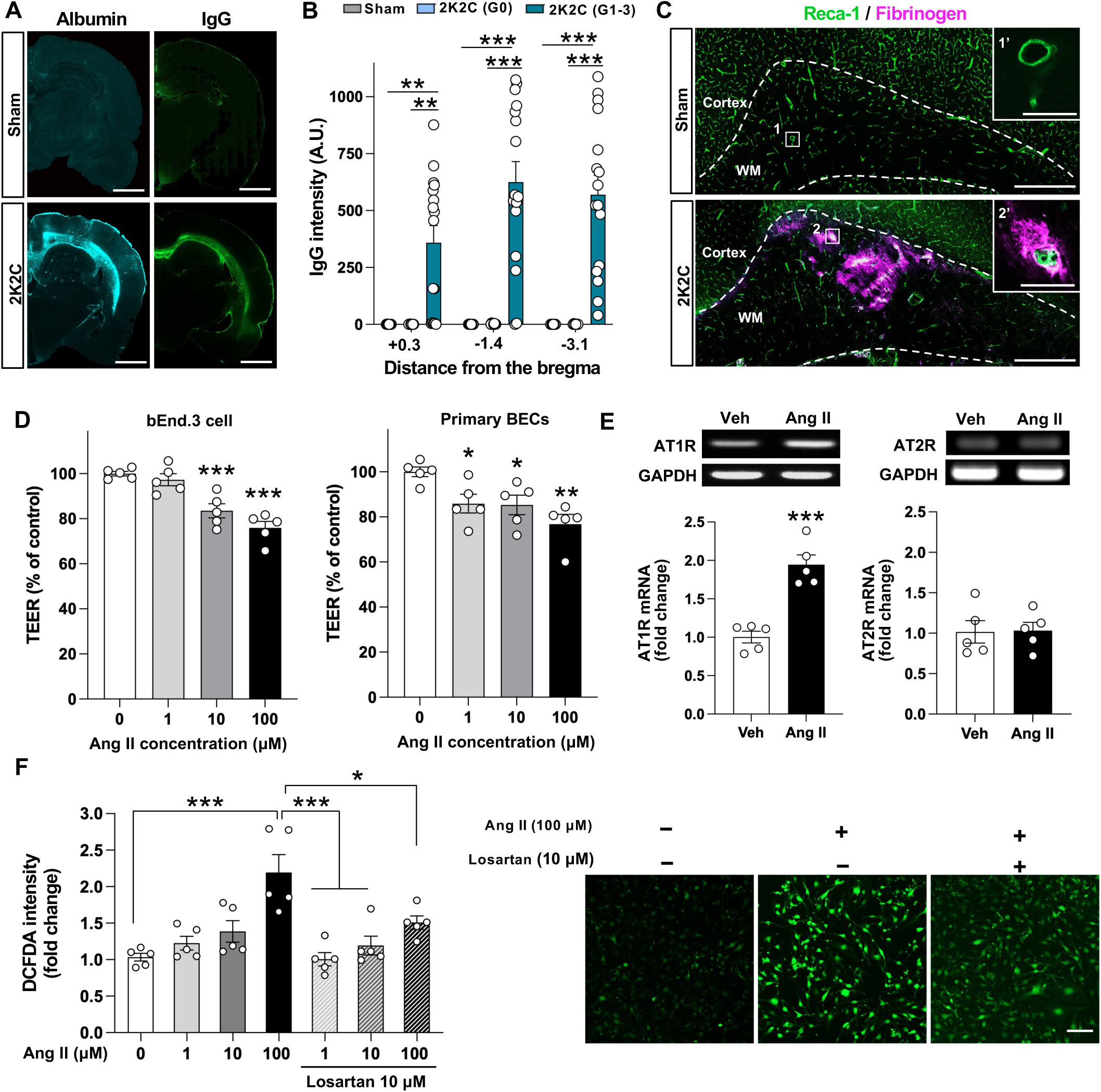
RVH-induced BBB disruption and implication of Ang II/AT1R signaling. **(A–C)** Assessment of BBB integrity in the white matter. **(A)** Representative images of coronal brain sections stained with antibodies against albumin and IgG in sham-operated and 2K2C rats. **(B)** Quantification graph comparing IgG immunoreactivity intensity in the white matter at 0.3 mm rostral, 1.4 mm, and 3.1 mm caudal to the bregma. A. U. indicates arbitrary units. Error bars represent mean ± SEM. ** and *** indicate P < 0.01 and P < 0.001, respectively. **(C)** Representative images of coronal brain sections double-stained with Reca-1 (green) for endothelial cells and fibrinogen (magenta) in sham-operated and 2K2C rats. Dashed lines demarcate the white matter. The boxed regions labeled with numbers are magnified in the right insets (1′, sham; 2′, 2K2C). n = 8 (sham), 8 (2K2C Grade 0), and 16 (2K2C Grade 1–3). Scale bars = 2 mm **(A)**, 500 μm and 50 μm (insets) **(C)**. **(D–F)** Assessment of Ang II-induced endothelial barrier dysfunction. **(D)** Quantification graphs comparing transendothelial electrical resistance (TEER) in bEnd.3 cells and primary brain endothelial cells (BECs) treated with the indicated concentrations of Ang II (μM) for 24 hours. n = 5 replicates per group. Each dot represents an independent culture replicate, and each biological replicate is an average of three technical replicates. **(E)** Representative RT-PCR images and quantification graphs comparing mRNA expression of AT1R and AT2R in bEnd.3 cells treated with vehicle (Veh) or Ang II. GAPDH was used as a loading control. n = 5 replicates per group. **(F)** Quantification graph comparing intracellular reactive oxygen species levels in bEnd.3 cells treated with Ang II in the presence or absence of losartan (10 μM). Error bars represent the mean ± SEM. *, **, and *** indicate *P* < 0.05, *P* < 0.01, and *P* < 0.001, respectively. n = 5 replicates per group. Each dot represents an independent culture replicate, and each biological replicate is an average of three technical replicates. Right panel: representative images of DCFDA fluorescence. Scale bar = 100 μm.

Ang II increases endothelial permeability, and AT1R activation can compromise BBB integrity.^29, 30^ We examined whether Ang II directly impairs barrier function in BEC monolayers by measuring TEER after 24 h of treatment with Ang II. Ang II reduced TEER in a dose-dependent manner in both b.End.3 and primary BECs isolated from adult mice (Fig. 5D), demonstrating that Ang II compromises endothelial barrier integrity. Because Ang II exerts its biological effects predominantly through AT1R,^31^ we examined its expression in bEnd.3 cells. RT-PCR confirmed that AT1R transcripts were present under basal conditions and were further upregulated following Ang II treatment (Fig. 5E), suggesting that this pathway is not only operative but also self-reinforcing in BECs. In contrast, Ang II treatment did not affect AT2R mRNA expression. As Ang II alters vascular function largely through ROS generation,^32, 33^ we confirmed that Ang II increased ROS production in a dose-dependent manner (Fig. 5F). To determine whether this response was AT1R-dependent, cells were co-treated with the AT1R blocker losartan (10 μM), which significantly attenuated the increase in ROS elicited by 100 μM Ang II (Fig. 5F). Together, these results indicate that Ang II drives ROS generation in BECs via AT1R, providing a plausible mechanism for the observed BBB impairment.

### Role of AT1R in WM pathology and neurological dysfunction in RVH rats

We next asked whether blocking AT1R signaling could prevent RVH-induced WM degeneration *in vivo*. Antagonizing peripheral AT1R would lower SBP and prevent WM injury in the first place. We reasoned that ICV delivery would allow us to target AT1R at the neurovascular interface while minimizing engagement of peripheral receptors (Fig. 6A). SBP rose progressively in both groups, with no significant difference between vehicle and losartan (Fig. 6B), verifying that ICV losartan did not exert appreciable systemic antihypertensive effects. There was a trend toward decreased mortality with losartan-treated RVH rats, 7 of 18 (38.9%) in aCSF-treated and 4 of 16 (25%) in losartan-treated animals, but this trend was not statistically significant by Chi-square test (*P* = 0.388).

**Figure 6.**
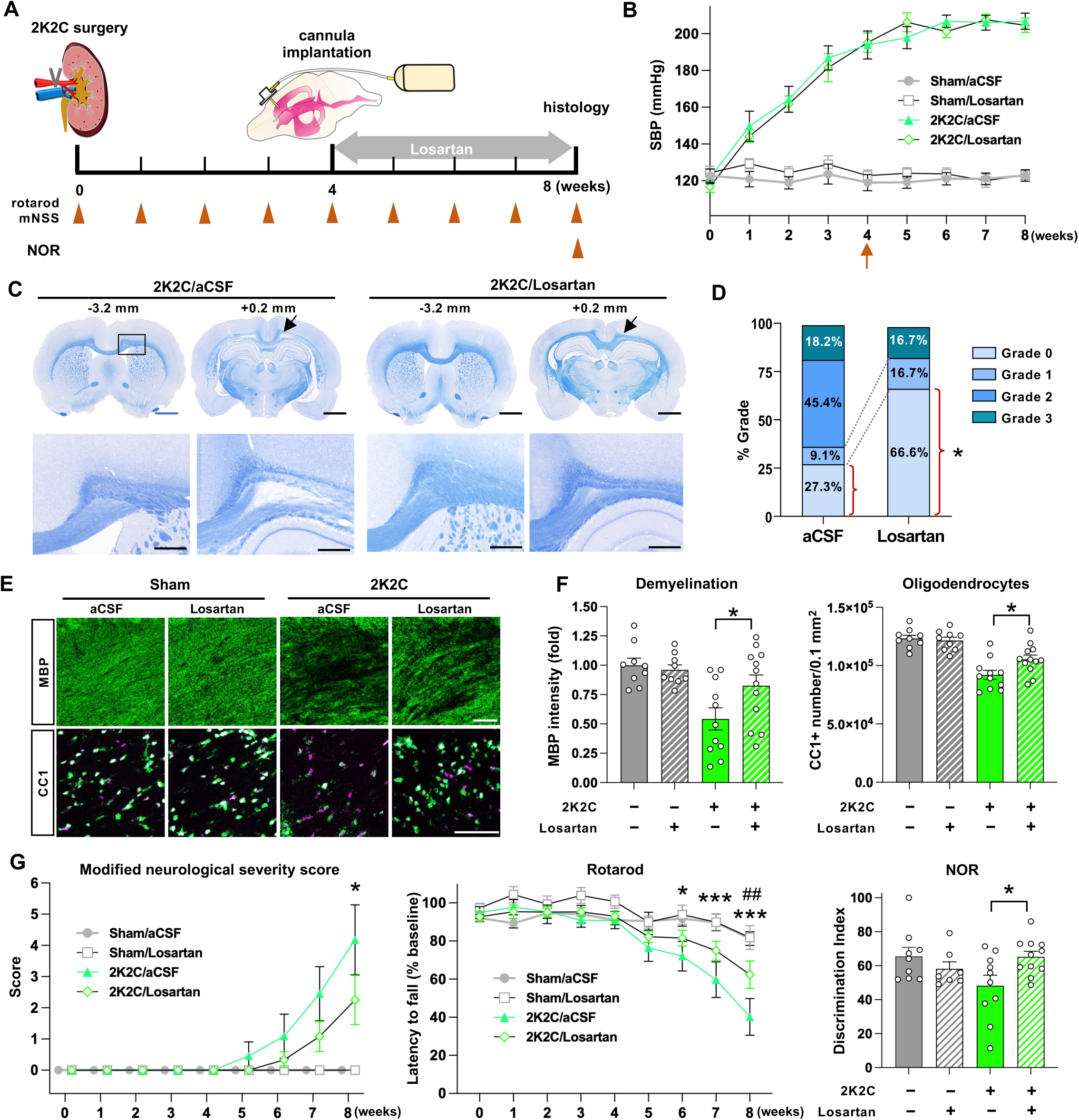
Intracerebroventricular losartan attenuates WM degeneration and neurological dysfunction in RVH rats. **(A)** Schematic diagram of the experimental design. Cannula implantation for intracerebroventricular (ICV) infusion of losartan or artificial cerebrospinal fluid (aCSF) was performed 4 weeks after 2K2C surgery, and infusion was maintained until 8 weeks. Arrowheads indicate the time points of behavioral assessment (rotarod, mNSS, and novel object recognition [NOR]). **(B)** Serial measurements of systolic blood pressure (SBP) in the four experimental groups. The arrow indicates the time of cannula implantation. **(C, D)** Assessment of WM lesion severity. **(C)** Representative images of coronal brain sections stained with Eriochrome Cyanine RC at 0.2 mm rostral and 3.2 mm caudal to the bregma in 2K2C rats infused with aCSF or losartan. The boxed region is magnified below. Arrows indicate areas of decreased myelin staining intensity in the corpus callosum and cingulum. Scale bars = 2 mm (upper panel) and 100 μm (lower panel). **(D)** Distribution of WM lesion severity grades in the aCSF- and losartan-infused groups. * indicates *P* < 0.05 by the chi-square test. Total n = 11 (aCSF) and 12 (losartan). **(E–G)** Assessment of demyelination and oligodendrocyte loss in the cingulum. **(E)** Representative images of coronal brain sections stained with antibodies against myelin basic protein (MBP) and APC-CC1 in sham-operated and 2K2C rats infused with aCSF or losartan. Scale bar = 50 μm. **(F)** Quantification graphs comparing the intensity of MBP immunoreactivity and the number of CC1-positive mature oligodendrocytes. * indicates *P* < 0.05 by two-way ANOVA followed by Bonferroni *post hoc* test. n = 9, 9, 11, and 12 for sham/aCSF, sham/losartan, 2K2C/aCSF, and 2K2C/losartan groups, respectively. **(G)** Neurological function assessment: Modified neurological severity score (mNSS) measured serially over 8 weeks; rotarod latency, expressed as a percentage of baseline; and discrimination index in the NOR test at 8 weeks. Error bars represent mean ± SEM. For mNSS and rotarod, * and *** indicate *P* < 0.05 and *P* < 0.001, respectively, versus the sham/aCSF group; ## indicates *P* < 0.01 between the 2K2C/aCSF and 2K2C/losartan groups, based on repeated measures two-way ANOVA followed by Bonferroni *post hoc* test. n = 9, 9, 11, and 12 for sham/aCSF, sham/losartan, 2K2C/aCSF, and 2K2C/losartan groups, respectively. For NOR test, * indicates *P* < 0.05 by two-way ANOVA followed by Bonferroni *post hoc* test. n = 9, 7, 10, and 11 for the sham/aCSF, sham/losartan, 2K2C/aCSF, and 2K2C/losartan groups, respectively (2 in sham/losartan, 1 in 2K2C/aCSF, and 1 in 2K2C/losartan groups were excluded due to incomplete tests)

Histological analysis of surviving animals showed that diffuse rarefaction of the CC or cingulum (Grade 2–3) was markedly less frequent after losartan infusion (Fig. 6C). Animals without apparent pathology comprised 3 of 11 (27.3%) in the aCSF group, comparable to untreated 2K2C rats (Fig. 2D), versus 8 of 12 (66.6%) after losartan (*P* < 0.05; Figure 6D). Grade 2 lesions were present in 5 of 11 (45.4%) aCSF-treated animals, but in none of those receiving losartan. Consistent with these findings, losartan partially preserved MBP immunoreactivity and the number of CC1-positive mature OLs in 2K2C animals (Fig. 6E, F). Two-way ANOVA revealed a statistically significant interaction between 2K2C surgery and losartan treatment for oligodendrocyte density (*P* < 0.05) but only a trend for MBP immunoreactivity (*P* = 0.0562). On *post hoc* testing, both measures were nonetheless significantly higher with losartan in 2K2C animals (*P* < 0.05; Fig. 6F).

We then tested if the histological benefits of losartan translated into functional improvement. mNSS scores rose beginning at 6 weeks in 2K2C animals, exceeding those of sham-operated animals (*P* < 0.05), although aCSF- and losartan-treated groups did not significantly differ (Fig. 6G). Rotarod performance likewise declined from 6 weeks; by 8 weeks, the latency to fall was longer with losartan than with aCSF among 2K2C animals (*P* < 0.01). In the NOR test, losartan almost entirely prevented the reduction in the discrimination index observed at 8 weeks in 2K2C animals, with a significant interaction between 2K2C surgery and drug treatment (*P* < 0.05) and a significantly higher discrimination index in the 2K2C/losartan than 2K2C/aCSF groups on *post hoc* testing (*P* < 0.05) (Figure 6G).

### *Post hoc* analyses of clinical cohorts to assess the association of antihypertensive class with WMHs

To further investigate the role of the renin-angiotensin system in WM ischemic injury in humans, we conducted *post hoc* analyses to assess the effect of antihypertensive class (ACEis, ARBs, BBs, CCBs, DIUs, and OTHs) on the progression of MRI-based WMHs. More detailed descriptions of the results are available in the Supplementary Results and Discussion. We started with two datasets from two independent clinical cohorts: the CHALLENGE study in South Korea (Fig. 7A)^22^ and the MSS2 in the UK (Fig. 7C)^23^. In the CHALLENGE study (n = 189), multivariable logistic regression analysis found that ARB use was consistently associated with higher odds of attenuated WMH progression across all outcomes: annual WMH progression ≤ 1st tertile (OR = 2.57, 95% CI [1.20-5.61]; *P* = 0.014), annual WMH progression ≤ cohort median (OR = 2.00, 95% CI [1.004-4.06]; *P* = 0.049), and annual WMH progression ≤ 0 (OR = 3.36, 95% CI [1.54-7.60]; *P* = 0.002) (Fig. 7B). In the MSS2 study (n = 191; Fig. 7C) multivariable logistic regression models revealed that no antihypertensive class, including ARBs and ACEis, was associated with changes in the odds of annual WMH progression for either the primary or secondary outcomes (Fig. 7D). An additional analysis combining users of ACEis and ARBs also did not reveal a significant association, indicating that the renin-angiotensin system does not influence WMH progression in the MSS2 UK cohort (Supplementary Fig. 4A). Because we were concerned that the short follow-up period (one year) may have contributed to the negative result in the MSS2 cohort, we analyzed the LBC1936 study (n = 465), another UK study with a 3-year follow-up. Multiple logistic regression did not reveal an association of any antihypertensive class with WMH progression(see Supplementary Results and Discussion for more detail).

**Figure 7.**
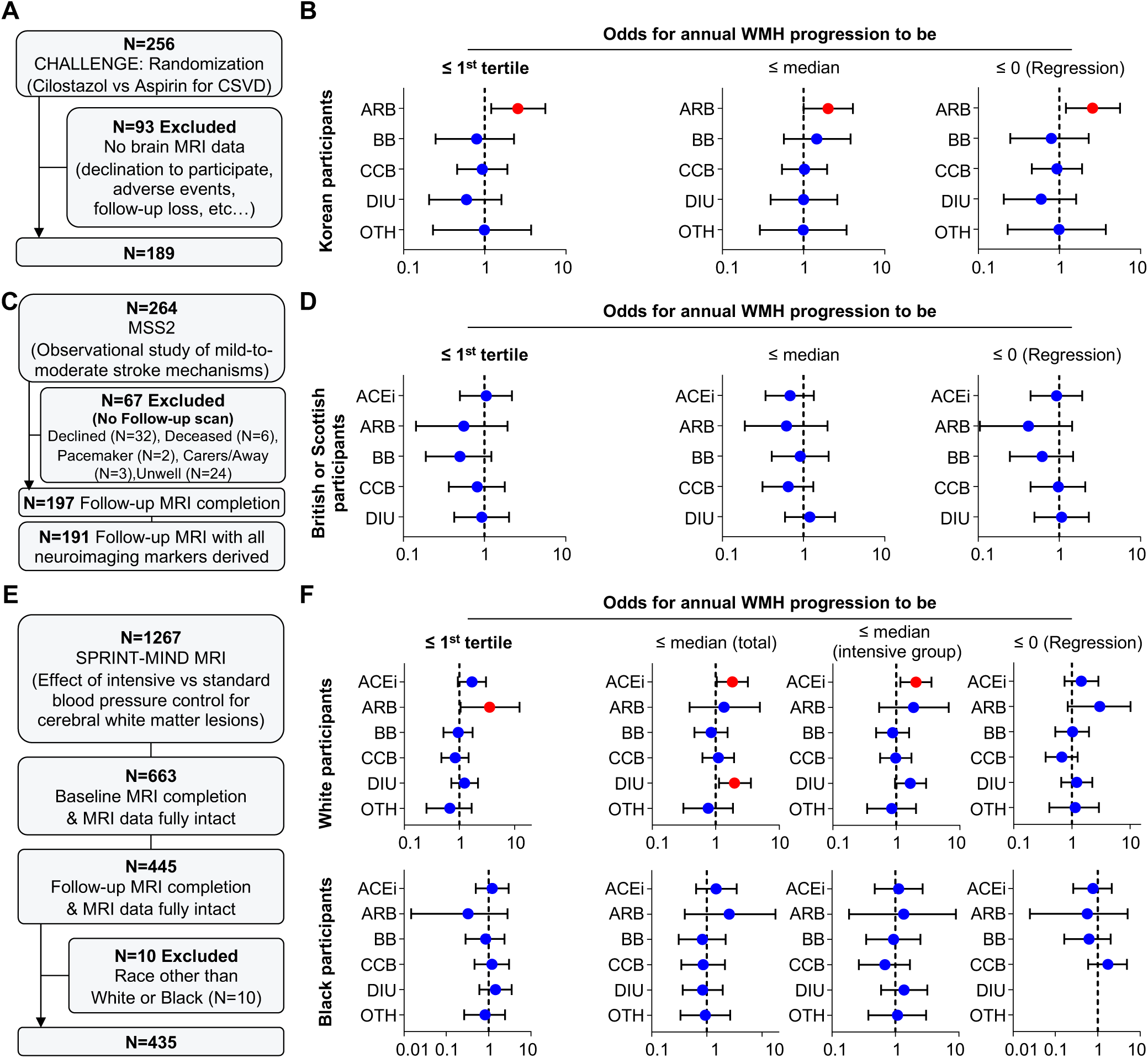
Effect of antihypertensive class on white matter hyperintensity progression in three human clinical cohorts. **(A)** Flowchart illustrating participant selection and follow-up in the CHALLENGE study. **(B)** Multivariable-adjusted odds ratios (ORs with 95% CIs) of antihypertensive classes for annualized WMH progression in the CHALLENGE cohort across three outcomes. **(C)** Flowchart of patient observation in the MSS2 cohort. **(D)** Multivariable-adjusted ORs (95% CIs) of antihypertensive classes for annualized WMH progression across three outcomes in the MSS2 cohort. **(E)** Flowchart illustrating participant selection and follow-up in the SPRINT-MIND MRI study. **(F)** Multivariable-adjusted ORs (95% CIs) for annualized WMH progression across four outcomes in the SPRINT-MIND MRI cohort.

The inconsistent results between the South Korean CHALLENGE study and the two UK studies, MSS2 and LBC1936, led us to hypothesize that ethnicity may influence how antihypertensive class affects WMH progression. Therefore, we revisited the SPRINT-MIND-MRI study in the US, in which a *post hoc* analysis reported that ACEis are most consistently associated with less WMH progression, independent of BP control and age.^34^ Curating the original dataset,^25^ we stratified participants by self-reported race into White (n = 300) and Black (n = 135) groups (Fig. 7E). In race-stratified multivariable logistic regression models, White participants receiving ARBs had significantly higher odds of annual WMH progression ≤ 1st tertile (OR = 3.52, 95% CI [1.05-12.29]; *P* = 0.042), whereas ACEi use was significantly associated with the secondary outcomes, higher odds of WMH progression ≤ overall median (OR = 1.84, 95% CI [1.05-3.24]; *P* = 0.035) and ≤ intensive-arm median (OR = 2.04, 95% CI [1.17-3.60]; *P* = 0.012) (Fig. 7F). Conversely, no antihypertensive class showed a significant protective effect against WMH progression among Black participants (Fig. 7F). Combining users of ACEis and ARBs did not alter the negative association among Black participants (Supplementary Fig. 4B). Collectively, these analyses suggest that ethnicity may play a role in the association between renin-angiotensin system inhibition and slower WMH progression.

## DISCUSSION

The results in this study replicated earlier reports showing that RVH induced by the 2K2C procedure manifests highly distinct WM lesions.^15–17^ Histopathological examination revealed clearly distinguishable foci of demyelination or diffuse regions with decreased myelin density, often referred to as rarefaction in the literature,^35, 36^ particularly in the cingulum and CC. Importantly, MBP and NF immunoreactivity and mature oligodendrocyte density in the CC were reduced by up to 50%, with effect sizes large enough to permit reliable quantification with conventional immunohistochemical measures. More striking were MRI findings that closely resembled human WMHs: T2 signal intensity progressively increased in WM regions, including the PVWM, the most common region in human WMHs. Our dMRI results were consistent with the structural data. The decreased FA and trends in other scalar metrics suggest more free diffusion of water and loss of microstructural integrity,^37^ and this matches the histological assessment.

To the authors’ knowledge, no other HT models have reported unequivocal pathological and MRI findings of this magnitude. Our previous study examined pathological alterations in the WM of spontaneously hypertensive rats (SHRs) using the same parameters as this study but found no changes in myelinated WM structures or mature OL numbers, even in SHRs subjected to vascular occlusion.^8^ Furthermore, stroke-prone SHRs (SHRSPs), which exhibit more severe HT and develop spontaneous stroke, did not show WMHs on MRI up to 10 months after birth.^38^ While some studies reported WM lesions in SHRs or SHRSPs, they did not present definitive evidence of WMHs on MRI,^39^ or lesion severity appeared far less than in our study.^39, 40^ We replicated the propensity of RVH to develop ischemic WM injury in a human study comparing MRI-based WMH burden with and without radiologically proven RAS. Notably, because all patients underwent CT to evaluate the potential cause of secondary hypertension, BP levels were comparable between groups with and without RAS. This finding suggests that alterations in the renin-angiotensin system may predict the severity of ischemic WM injury independently of BP levels. In accordance with our analysis, polymorphisms in genes involved in the renin-angiotensin system were significantly associated with the presence of WMHs after controlling for HT.^41^

Our further investigations, integrating *in vitro*, *in vivo* animal, and human clinical data, established the potential role of Ang II and AT1R signaling in BBB impairment and ischemic WM injury. Impaired BBB barrier function is a core mechanism driving CSVD progression.^26^ Our *in vitro* study demonstrated that Ang II induces ROS generation and directly impairs barrier function via AT1R expressed in BECs. Importantly, ICV infusion of losartan substantially reduced WM injury and significantly ameliorated neurological dysfunction in RVH rats, supporting the notion that Ang II and AT1R may be critically implicated in the evolution of WM ischemic injury. One caveat is that AT1R is expressed in other brain cell types, such as dopaminergic neurons and microglial cells.^42, 43^ More studies are needed to clarify the potential influence of AT1R inhibition on glial cells involved in WM ischemic injury. Our *post hoc* analyses of the clinical cohorts support the animal findings by showing that renin-angiotensin system inhibitors are associated with slower WMH progression, although this association was not uniform across cohorts. In the CHALLENGE cohort, ARB use was consistently associated with slower WMH progression across primary and secondary outcomes. A comparable association was observed among White participants in SPRINT-MIND-MRI, in whom ARBs and ACEis were associated with the primary and secondary outcomes, respectively. The association was not replicated in the MSS2 and LBC1936 studies, both predominantly White Scottish cohorts, suggesting that the effect is not consistently detectable across all White populations. However, factors other than ethnicity may have contributed to the disparities in results across different cohorts (see Supplementary Results and Discussion for more detail). Among Black participants in SPRINT-MIND-MRI, renin-angiotensin system inhibitors were not associated with slower WMH progression. One possible contributor to these differences is genetic variation governing AT1R expression in BECs. The A1166C single-nucleotide polymorphism in the 3′ untranslated region of the AT1R transcript increases receptor abundance,^44^ and its allelic frequency is lowest in populations of African ancestry.^45^ We speculate that a lower burden of this variant reduces the contribution of AT1R signaling to WM injury, and correspondingly the benefit derived from blocking it, although genotypes were not available in these cohorts.

Although the RVH rat model in this study showed distinct and unequivocal WM lesions on MRI and histopathology, a critical limitation as a disease model was that approximately one-third of RVH rats did not develop WM lesions by 9 weeks after 2K2C surgery. Previous studies have also reported inconsistent incidence and severity of WM lesions.^15, 16^ The lower mean SBP in animals with pathology grade 0 may impose less hemodynamic stress on endothelial cells, thereby preserving microcirculation within the WM.^46^ However, ICV losartan substantially attenuated WM injury, even though BP remained comparable to the vehicle condition. When SHRs were given either the vasodilator hydralazine or losartan, losartan, but not hydralazine, prevented BBB disruption, even though BP reductions were similar for both drugs.^47^ Furthermore, the presence of RAS in our human cohort predicted a higher WMH burden without affecting SBP, suggesting that BP itself is not a strong determinant of the extent of BBB impairment or WM injury. Given the genetic heterogeneity of the outbred SD strain, resilience-related genetic factors may also contribute to the inconsistent manifestation of WM lesions in the RVH model. Lin et al. addressed this inconsistency by prolonging RVH duration and combining occlusion of the common carotid arteries.^16^ More studies are needed to optimize this model, as combining either approach would increase mortality risk.

Our results suggest that drugs that inhibit the renin-angiotensin system, specifically ARBs, may offer therapeutic benefits for WM ischemic lesions and associated cognitive decline. Previous studies suggested that ARBs may reduce the risk of dementia and stroke. ARB medication for older male hypertensive patients significantly reduced the hazard rate of incident dementia and, in patients with pre-existing dementia, was associated with a lower risk of nursing home admission or death.^48^ The beneficial effects of ARBs are broadly attributed to pleiotropic neuroprotective mechanisms, including antioxidant and anti-inflammatory effects.^49^ AT1R is expressed in other cells at the neurovascular interface, such as pericytes and border-associated macrophages.^50, 51^ AT1R in these cells is also implicated in BBB integrity and microcirculation regulation. Therefore, ARBs with higher BBB penetrance may be more advantageous in preventing ischemic WM injury. Our clinical study suggests that the beneficial effects of ARBs may depend on ethnicity. Future prospective RCTs examining the effects of ARBs on the progression of WM ischemic degeneration and on cognitive and cardiovascular health should therefore include ethnically diverse participants.

## Acknowledgments

We thank the Institute for Basic Science (IBS) Center for Neuroscience Imaging Research (IBS-R015-D1-2016-a00) for providing animal scanner time and professional technical support. We thank Prof. Yuhua Fan of Sun Yat-sen University in Guangzhou, China, for generously providing silver clips.

## Data availability

The data supporting the findings of this study are available from the corresponding authors upon reasonable request. Data from SPRINT-MIND-MRI are publicly available through the NHLBI BioLINCC repository (https://biolincc.nhlbi.nih.gov/studies/sprint/). Data from the CHALLENGE, MSS2, LBC1936 studies are available from the original investigators. The interpretations presented here are those of the authors and do not necessarily reflect the views of the original study investigators or sponsors.

## Reference

1. Pantoni L. Cerebral small vessel disease: From pathogenesis and clinical characteristics to therapeutic challenges. Lancet neurology. 2010;9:689–701

2. Dichgans M, Leys D. Vascular cognitive impairment. Circ Res. 2017;120:573–591

3. Gorelick PB, Scuteri A, Black SE, Decarli C, Greenberg SM, Iadecola C, et al. Vascular contributions to cognitive impairment and dementia: A statement for healthcare professionals from the american heart association/american stroke association. Stroke; a journal of cerebral circulation. 2011;42:2672–2713

4. Attems J, Jellinger KA. The overlap between vascular disease and alzheimer’s disease--lessons from pathology. BMC Med. 2014;12:206

5. Liu Y, Chan DK, Crawford JD, Sachdev PS, Braidy N. The contribution of cerebral vascular neuropathology to mild stage of alzheimer’s dementia using the nacc database. Curr Alzheimer Res. 2020;17:1167–1176

6. Strickland S. Blood will out: Vascular contributions to alzheimer’s disease. The Journal of clinical investigation. 2018;128:556–563

7. Marin MA, Carmichael ST. Stroke in cns white matter: Models and mechanisms. Neuroscience letters. 2018;684:193–199

8. Choi JY, Cui Y, Kim BG. Interaction between hypertension and cerebral hypoperfusion in the development of cognitive dysfunction and white matter pathology in rats. Neuroscience. 2015;303:115–125

9. Cui Y, Jin X, Choi JY, Kim BG. Modeling subcortical ischemic white matter injury in rodents: Unmet need for a breakthrough in translational research. Neural Regen Res. 2021;16:638–642

10. Reimer MM, McQueen J, Searcy L, Scullion G, Zonta B, Desmazieres A, et al. Rapid disruption of axon-glial integrity in response to mild cerebral hypoperfusion. J Neurosci. 2011;31:18185–18194

11. Füchtemeier M, Brinckmann MP, Foddis M, Kunz A, Po C, Curato C, et al. Vascular change and opposing effects of the angiotensin type 2 receptor in a mouse model of vascular cognitive impairment. Journal of cerebral blood flow and metabolism : official journal of the International Society of Cerebral Blood Flow and Metabolism. 2015;35:476–484

12. Hall GR, Boehm-Sturm P, Dirnagl U, Finke C, Foddis M, Harms C, et al. Long-term connectome analysis reveals reshaping of visual, spatial networks in a model with vascular dementia features. Stroke; a journal of cerebral circulation. 2022;53:1735–1745

13. Masumura M, Hata R, Nagai Y, Sawada T. Oligodendroglial cell death with DNA fragmentation in the white matter under chronic cerebral hypoperfusion: Comparison between normotensive and spontaneously hypertensive rats. Neuroscience Research. 2001;39:401–412

14. Jalal FY, Yang Y, Thompson J, Lopez AC, Rosenberg GA. Myelin loss associated with neuroinflammation in hypertensive rats. Stroke; a journal of cerebral circulation. 2012;43:1115–1122

15. Fan Y, Lan L, Zheng L, Ji X, Lin J, Zeng J, et al. Spontaneous white matter lesion in brain of stroke-prone renovascular hypertensive rats: A study from mri, pathology and behavior. Metabolic Brain Disease. 2015;30:1479–1486

16. Lin J, Lan L, Wang D, Qiu B, Fan Y. Cerebral venous collagen remodeling in a modified white matter lesions animal model. Neuroscience. 2017;367:72–84

17. Lan LF, Zheng L, Yang X, Ji XT, Fan YH, Zeng JS. Peroxisome proliferator-activated receptor-γ agonist pioglitazone ameliorates white matter lesion and cognitive impairment in hypertensive rats. CNS Neurosci Ther. 2015;21:410–416

18. Zeng J, Zhang Y, Mo J, Su Z, Huang R. Two-kidney, two clip renovascular hypertensive rats can be used as stroke-prone rats. Stroke; a journal of cerebral circulation. 1998;29:1708–1713; discussion 1713-1704

19. Daugherty A, Rateri D, Hong L, Balakrishnan A. Measuring blood pressure in mice using volume pressure recording, a tail-cuff method. Journal of visualized experiments : JoVE. 2009

20. Schwarz AJ, Danckaert A, Reese T, Gozzi A, Paxinos G, Watson C, et al. A stereotaxic mri template set for the rat brain with tissue class distribution maps and co-registered anatomical atlas: Application to pharmacological mri. NeuroImage. 2006;32:538–550

21. Tierney TM, Alexander NA, Ashburner J, Avila NL, Balbastre Y, Barnes G, et al. Spm 25: Open source neuroimaging analysis software. Journal of Open Source Software. 2025;10

22. Kim BC, Youn YC, Jeong JH, Han HJ, Kim JH, Lee J-H, et al. Cilostazol versus aspirin on white matter changes in cerebral small vessel disease: A randomized controlled trial. Stroke; a journal of cerebral circulation. 2022;53:698–709

23. Valdés Hernández MdC, Armitage PA, Thrippleton MJ, Chappell F, Sandeman E, Muñoz Maniega S, et al. Rationale, design and methodology of the image analysis protocol for studies of patients with cerebral small vessel disease and mild stroke. Brain and Behavior. 2015;5

24. Deary IJ, Gow AJ, Taylor MD, Corley J, Brett C, Wilson V, et al. The lothian birth cohort 1936: A study to examine influences on cognitive ageing from age 11 to age 70 and beyond. BMC Geriatrics. 2007;7

25. Nasrallah IM, Pajewski NM, Auchus AP, Chelune G, Cheung AK, Cleveland ML, et al. Association of intensive vs standard blood pressure control with cerebral white matter lesions. JAMA. 2019;322:524

26. Wardlaw JM, Makin SJ, Valdés Hernández MC, Armitage PA, Heye AK, Chappell FM, et al. Blood-brain barrier failure as a core mechanism in cerebral small vessel disease and dementia: Evidence from a cohort study. Alzheimer’s & dementia : the journal of the Alzheimer’s Association. 2017;13:634–643

27. Fan Y, Yang X, Tao Y, Lan L, Zheng L, Sun J. Tight junction disruption of blood-brain barrier in white matter lesions in chronic hypertensive rats. Neuroreport. 2015;26:1039–1043

28. Petersen MA, Ryu JK, Akassoglou K. Fibrinogen in neurological diseases: Mechanisms, imaging and therapeutics. Nature reviews. Neuroscience. 2018;19:283–301

29. Fleegal-DeMotta MA, Doghu S, Banks WA. Angiotensin ii modulates bbb permeability via activation of the at(1) receptor in brain endothelial cells. Journal of cerebral blood flow and metabolism : official journal of the International Society of Cerebral Blood Flow and Metabolism. 2009;29:640–647

30. Guo S, Som AT, Arai K, Lo EH. Effects of angiotensin-ii on brain endothelial cell permeability via pparalpha regulation of para- and trans-cellular pathways. Brain research. 2019;1722:146353

31. Saito Y, Berk BC. Angiotensin ii-mediated signal transduction pathways. Curr Hypertens Rep. 2002;4:167–171

32. Girouard H, Park L, Anrather J, Zhou P, Iadecola C. Angiotensin ii attenuates endothelium-dependent responses in the cerebral microcirculation through nox-2-derived radicals. Arteriosclerosis, thrombosis, and vascular biology. 2006;26:826–832

33. Manuneedhi Cholan P, Cartland SP, Dang L, Rayner BS, Patel S, Thomas SR, et al. Trail protects against endothelial dysfunction in vivo and inhibits angiotensin-ii-induced oxidative stress in vascular endothelial cells in vitro. Free radical biology & medicine. 2018;126:341–349

34. Goldstein ED, Wolcott Z, Garg G, Navarro K, Delic A, Yaghi S, et al. Effect of antihypertensives by class on cerebral small vessel disease: A post hoc analysis of sprint-mind. Stroke; a journal of cerebral circulation. 2022;53:2435–2440

35. Wakita H, Tomimoto H, Akiguchi I, Matsuo A, Lin JX, Ihara M, et al. Axonal damage and demyelination in the white matter after chronic cerebral hypoperfusion in the rat. Brain research. 2002;924:63–70

36. Shibata M, Ohtani R, Ihara M, Tomimoto H. White matter lesions and glial activation in a novel mouse model of chronic cerebral hypoperfusion. Stroke; a journal of cerebral circulation. 2004;35:2598–2603

37. Jones DK, Knösche TR, Turner R. White matter integrity, fiber count, and other fallacies: The do’s and don’ts of diffusion mri. NeuroImage. 2013;73:239–254

38. Brittain JF, McCabe C, Khatun H, Kaushal N, Bridges LR, Holmes WM, et al. An mri-histological study of white matter in stroke-free shrsp. Journal of Cerebral Blood Flow & Metabolism. 2013;33:760–763

39. Lin JX, Tomimoto H, Akiguchi I, Wakita H, Shibasaki H, Horie R. White matter lesions and alteration of vascular cell composition in the brain of spontaneously hypertensive rats. Neuroreport. 2001;12:1835–1839

40. Kaiser D, Weise G, Moller K, Scheibe J, Posel C, Baasch S, et al. Spontaneous white matter damage, cognitive decline and neuroinflammation in middle-aged hypertensive rats: An animal model of early-stage cerebral small vessel disease. Acta Neuropathol Commun. 2014;2:169

41. Sierra C, Coca A, Gómez-Angelats E, Poch E, Sobrino J, de la Sierra A. Renin-angiotensin system genetic polymorphisms and cerebral white matter lesions in essential hypertension. Hypertension. 2002;39:343–347

42. Zhang W, Jiao B, Yu S, Zhang K, Sun J, Liu B, et al. Spinal at1r contributes to neuroinflammation and neuropathic pain via nox2-dependent redox signaling in microglia. Free Radical Biology and Medicine. 2025;227:143–156

43. Daynac M, Mouilleau V, Menendez A, Liu Y, Yang X, Shen Y, et al. A druggable link between the angiotensin receptor 1 and α-synuclein pathology revealed in scalable human cellular models of parkinson’s disease. Molecular Neurodegeneration Advances. 2026;2:32

44. Baudin B. Angiotensin ii receptor polymorphisms in hypertension. Pharmacogenomic considerations. Pharmacogenomics. 2002;3:65–73

45. Mottl AK, Shoham DA, North KE. Angiotensin ii type 1 receptor polymorphisms and susceptibility to hypertension: A huge review. Genet Med. 2008;10:560–574

46. Joutel A, Monet-Leprêtre M, Gosele C, Baron-Menguy C, Hammes A, Schmidt S, et al. Cerebrovascular dysfunction and microcirculation rarefaction precede white matter lesions in a mouse genetic model of cerebral ischemic small vessel disease. Journal of Clinical Investigation. 2010;120:433–445

47. Biancardi VC, Stern JE. Compromised blood-brain barrier permeability: Novel mechanism by which circulating angiotensin ii signals to sympathoexcitatory centres during hypertension. J Physiol. 2016;594:1591–1600

48. Li NC, Lee A, Whitmer RA, Kivipelto M, Lawler E, Kazis LE, et al. Use of angiotensin receptor blockers and risk of dementia in a predominantly male population: Prospective cohort analysis. BMJ. 2010;340:b5465–b5465

49. Villapol S, Saavedra JM. Neuroprotective effects of angiotensin receptor blockers. Am J Hypertens. 2015;28:289–299

50. Kawamura H, Kobayashi M, Li Q, Yamanishi S, Katsumura K, Minami M, et al. Effects of angiotensin ii on the pericyte-containing microvasculature of the rat retina. J Physiol. 2004;561:671–683

51. Li T, Zhang J, Song H, Zhang R, Fan F, Huang Z, et al. Border-associated macrophages: An emerging perspective from physiological basis and multi-disease roles to the mechanism of vascular cognitive impairment and dementia. Journal of neuroinflammation. 2025;22:302

